# Novel *KITLG/SCF* regulatory variants are associated with lung function in African American children with asthma

**DOI:** 10.1101/2020.02.20.20019588

**Authors:** Angel CY Mak, Satria Sajuthi, Jaehyun Joo, Shujie Xiao, Patrick M Sleiman, Marquitta J White, Eunice Y Lee, Benjamin Saef, Donglei Hu, Hongsheng Gui, Kevin L Keys, Fred Lurmann, Deepti Jain, Gonçalo Abecasis, Hyun Min Kang, Deborah A. Nickerson, Soren Germer, Michael C Zody, Lara Winterkorn, Catherine Reeves, Scott Huntsman, Celeste Eng, Sandra Salazar, Sam S Oh, Frank D Gilliland, Zhanghua Chen, Rajesh Kumar, Fernando D Martínez, Ann Chen Wu, Elad Ziv, Hakon Hakonarson, Blanca E Himes, L Keoki Williams, Max A Seibold, Esteban G. Burchard

## Abstract

Baseline lung function, quantified as forced expiratory volume in the first second of exhalation (FEV_1_), is a standard diagnostic criterion used by clinicians to identify and classify lung diseases. Using whole genome sequencing data from the National Heart, Lung, and Blood Institute TOPMed project, we identified a novel genetic association with FEV_1_ on chromosome 12 in 867 African American children with asthma (p = 1.26 × 10^−8^, β = 0.302). Conditional analysis within 1 Mb of the tag signal (rs73429450) yielded one major and two other weaker independent signals within this peak. We explored statistical and functional evidence for all variants in linkage disequilibrium with the three independent signals and yielded 9 variants as the most likely candidates responsible for the association with FEV_1_. Hi-C data and eQTL analysis demonstrated that these variants physically interacted with *KITLG (aka SCF)* and their minor alleles were associated with increased expression of *KITLG* gene in nasal epithelial cells. Gene-by-air-pollution interaction analysis found that the candidate variant rs58475486 interacted with past-year SO_2_ exposure (p = 0.003, β = 0.32). This study identified a novel protective genetic association with FEV_1_, possibly mediated through *KITLG*, in African American children with asthma.

## INTRODUCTION

Asthma, a chronic pulmonary condition characterized by reversible airway obstruction, is one of the hallmark diseases of childhood in the United States (World Health Organization 2017). Asthma is also the most disparate common disease in the pediatric clinic, with significant variation in prevalence, morbidity, and mortality among U.S. racial/ethnic groups (Oh *et al*. 2016). Specifically, African American children carry a higher asthma disease burden compared to their European American counterparts (Akinbami *et al*. 2014; Akinbami 2015). Forced expiratory volume in the first second (FEV_1_), a measurement of lung function, is a vital clinical trait used by physicians to assess overall lung health and diagnose pulmonary diseases such as asthma (Johnson and Theurer. 2014). We have previously shown that genetic ancestry plays an important role in FEV_1_ variation and that African Americans have lower FEV_1_ compared to European Americans regardless of asthma status (Kumar *et al*. 2010; Pino-Yanes *et al*. 2015). The disparity in lung function between populations may explain disparities in asthma disease burden. Understanding the factors that influence FEV_1_ variation among individuals with asthma could lead to improved patient care and therapeutic interventions.

Twin and family-based studies estimate that the heritability of FEV_1_ ranged from 26% to 81%, supporting the combined contribution by genetic and environmental factors in FEV_1_ variation (Chatterjee and Das. 1995; Chen *et al*. 1996; Hukkinen *et al*. 2011; Palmer *et al*. 2001; Sillanpaa *et al*. 2017; Tian *et al*. 2017; Yamada *et al*. 2015). Genome-wide association studies (GWAS) of FEV_1_, including among individuals with asthma, have identified many variants that contribute to lung function (Li *et al*. 2013; Liao *et al*. 2014; Repapi *et al*. 2010; Soler Artigas *et al*. 2011; Soler Artigas *et al*. 2015; Wain *et al*. 2017). A search in NHGRI-EBI GWAS Catalog (version e98_r2020-03-08) on baseline lung function (FEV_1_) alone revealed 349 associations (Buniello *et al*. 2019). Most of these previous GWAS, however, were performed in adult populations of European descent, and their results may not generalize across populations or across the life span of an individual (Carlson *et al*. 2013; Martin, A. R. *et al*. 2017; Wojcik *et al*. 2019). Previous GWAS results are also limited due to their reliance on genotyping arrays. In particular, variation in non-coding regions of the genome is not adequately covered by many genotyping arrays because they were not designed to account for the population-specific genetic variability of all populations (Kim, M. S. *et al*. 2018; Zhang and Lupski. 2015). Whole genome sequencing (WGS) is a newer technology that captures nearly all common variation from coding and non-coding regions of the genome and is unencumbered by genotype array design constraints and differences in linkage disequilibrium patterns among populations. To date, no large-scale WGS studies of lung function have been performed in African American children with asthma (Martin *et al*. 2017).

In addition to genetics, FEV_1_ is a complex trait that is significantly influenced by both genetic variation and environmental factors, such as air pollution (Chatterjee and Das. 1995; Hukkinen *et al*. 2011; Palmer *et al*. 2001; Sillanpaa *et al*. 2017; Tian *et al*. 2017; Yamada *et al*. 2015). Exposure to ambient air pollution has been consistently associated with poor respiratory outcomes, including reduced FEV_1_ (Barraza-Villarreal *et al*. 2008; Brunekreef and Holgate. 2002; Ierodiakonou *et al*. 2016; Wise 2019). We previously showed that exposure to sulfur dioxide (SO_2_), an air pollutant emitted by the burning of fossil fuels, is significantly associated with reduced FEV_1_ in African American children with asthma in the SAGE II study (Neophytou *et al*. 2016). Because the genetic variants associated with FEV_1_ thus far do not account for the majority of its estimated heritability, considering gene-environment (GxE) interactions, specifically gene-by-air-pollution, may improve our understanding of lung function genetics (Moore 2005; Moore and Williams. 2009). Here, we performed a genome-wide association analysis using WGS data to identify common genetic variants associated with FEV_1_ in African American children with asthma in SAGE II and investigated the effect of GxE (SO_2_) interactions on FEV_1_ associations.

## METHODS

### Study population

This study examined African American children between 8-21 years of age with physician-diagnosed asthma from the Study of African Americans, Asthma, Genes & Environments (SAGE II). All SAGE II participants were recruited from the San Francisco Bay Area. The inclusion and exclusion are previously described in detailed (Oh *et al*. 2012; White *et al*. 2016). Briefly, participants were eligible if they were 8-21 years of age and self-identified as African American and had four African American grandparents. Study exclusion criteria included the following: 1) any smoking within one year of the recruitment date; 2) 10 or more pack-years of smoking; 3) pregnancy in the third trimester; 4) history of lung diseases other than asthma (for cases) or chronic illness (for cases and controls). Baseline lung function defined as forced expiratory volume in the first second (FEV_1_) was measured by spirometry prior to administering albuterol as previously described (Oh *et al*. 2012).

### TOPMed whole genome sequencing data

SAGE II DNA samples were sequenced as part of the Trans-Omics for Precision Medicine (TOPMed) whole genome sequencing (WGS) program (Taliun *et al*. 2019). WGS was performed at the New York Genome Center and Northwest Genomics Center on a HiSeq X system (Illumina, San Diego, CA) using a paired-end read length of 150 base pairs (bp), with a minimum of 30x mean genome coverage. DNA sample handling, quality control, library construction, clustering and sequencing, read processing and sequence data quality control are described in detail in the TOPMed website (TOPMed 2019). Variant calls were obtained from TOPMed data freeze 8 VCF files corresponding to the GRCh38 assembly. Variants with a minimal read depth of 10 (DP10) were used for analysis unless otherwise stated.

### Genetic principal components, global ancestry, and kinship estimation

Genetic principal components (PCs), global ancestry, and kinship estimation on genetic relatedness were computed using biallelic single nucleotide polymorphisms (SNPs) with a PASS flag from TOPMed freeze 8 DP10 data. PCs and kinship estimates were computed using the PC-Relate function from the GENESIS R package (Conomos *et al*. 2015; Conomos *et al*. 2016) using a workflow available from the Summer Institute in Statistical Genetics Module 17 course website (Summer Institute in Statistical Genetics 2019). African global ancestry was computed using the ADMIXTURE package (Alexander *et al*. 2009) in supervised mode using European (CEU), African (YRI) and Native American (NAM) reference panels as previously described (Mak, A. C. Y. *et al*. 2018).

### FEV_1_ GWAS

Non-normality of the distribution of FEV_1_ values was tested with the Shapiro-Wilk test in R using the shapiro.test function. Since FEV_1_ was not normally distributed (p = 1.41 × 10^−8^ for FEV_1_ and p = 1.05 × 10^−8^ for log_10_ FEV_1_), FEV_1_ was regressed on all covariates (age, sex, height, controller medications, sequencing centers, and the first 5 genetic PCs) and the residuals were inverse-normalized. These inverse-normalized residuals (FEV_1_.res.rnorm) were the main outcome of the discovery GWAS. The controller medication covariate included the use of inhaled corticosteroids (ICS), long-acting beta-agonists (LABA), leukotriene inhibitors and/or an ICS/LABA combo in the 2 weeks prior to the recruitment date.

Genome-wide single variant analysis was performed on the ENCORE server (https://github.com/statgen/encore) using the linear Wald test (q.linear) originally implemented in EPACTS (https://genome.sph.umich.edu/wiki/EPACTS) and TOPMed freeze 8 data (DP0 PASS) with a MAF filter of 0.1%. All pairwise relationships with degree 3 or more relatedness (kinship values > 0.044) were identified, and one participant of the related pair was subsequently chosen at random and removed prior to analysis. All covariates used to obtain FEV_1_.res.rnorm were also included as covariates in the GWAS as recommended in a recent publication (Sofer *et al*. 2019). The association analysis was repeated using untransformed FEV_1_ and FEV_1_ percent predicted (FEV_1_.perc.predicted). FEV_1_ percent predicted was defined as the percentage of measured FEV_1_ relative to predicted FEV_1_ estimated by the Hankinson lung function prediction equation for African Americans (Hankinson *et al*. 1999). A secondary analysis that included smoking-related covariates (smoking status and number of smokers in the family) was performed in PLINK 1.9 (version 1p9_2019_0304_dev) (Chang *et al*. 2015; Purcell and Chang. 2013). To study whether association with FEV_1_ is specific to SAGE II participants with asthma, we repeated the association analysis adjusting for age, sex, height and the first 5 genetic PCs in SAGE II participants without asthma on the ENCORE server. All of these participants were sequenced in the same center. Regional association results were plotted using LocusZoom 1.4 (Pruim *et al*. 2010) with a 500 kilobase (Kb) flanking region. Linkage disequilibrium (R^2^) was estimated in PLINK 1.9. LD plot was generated using recoded genotype files (plink --recode 12) in Haploview (Barrett *et al*. 2005).

The function effectiveSize in the R package CODA was used to estimate the actual effective number of independent tests and CODA-adjusted statistical and suggestive significance p-value thresholds were defined as 0.05 and 1 divided by the effective number of tests, respectively (Duggal *et al*. 2008). We compared the CODA-adjusted statistical significance threshold and the widely used 5 × 10^−8^ GWAS genome-wide significance threshold (Pe’er *et al*. 2008) and selected the more stringent threshold for genome-wide significance.

The following WGS quality control steps were applied to all reported variants from ENCORE to ensure WGS variant quality: (1) The variant had VCF FILTER = PASS; (2) Variant quality was confirmed via manual inspection on the BRAVO server based on TOPMed freeze 5 data (University of Michigan and NHLBI TOPMed. 2018); (3) Variants were reanalyzed with linear regression using PLINK 1.9 by applying the arguments --mac 5 --geno 0.1 --hwe 0.0001 using TOPMed freeze 8 DP10 PASS data.

To determine if the rs73429450 association with FEV_1_ was only identifiable using whole genome sequencing data, we repeated the linear regression association analysis on signals that passed the genome-wide significance threshold using PLINK 1.9 and genotype data generated with Axiom Genome-Wide LAT 1 array (Affymetrix, Santa Clara, CA, dbGaP phs000921.v1.p1). These array genotype data were imputed into the following reference panels: 1000 Genomes phase 3 version 5, Haplotype Reference Consortium (HRC) r1.1, the Consortium on Asthma among African-ancestry Populations in the Americas (CAAPA) and the TOPMed phase 5 panels on the Michigan Imputation Server (Das *et al*. 2016). It should be noted that 500 SAGE II subjects were part of the TOPMed freeze 5 reference panel.

A total of 349 GWAS FEV_1_-associated entries were retrieved from the NHGRI-EBI GWAS Catalog version 1.0.2-associations_e98_r2020-03-08 (Buniello *et al*. 2019) using the trait names “Lung function (FEV_1_)”, “FEV1”, “Lung function (forced expiratory volume in 1 second)” or “Prebronchodilator FEV1”. After adding 100 Kb flanking regions to each of the 349 entries, a total of 230 non-overlapping region were obtained. To look up whether we replicated previously GWAS loci while control for multiple testing penalties, we only used 279,495 common variants (MAF >= 0.01) that overlapped with the 230 regions. The 279,495 common variants is equivalent to 17,755 effective test based on CODA and 5.63 × 10^−5^ (1/17,755) was used as suggestive p-value threshold for replication.

### Conditional analysis

Conditional analysis was performed to identify all independent signals in a GWAS peak using PLINK 1.9. All TOPMed freeze 8 DP10 variants within 1 megabase (Mb) of the tag association signal and with association p-value of 1 × 10^−4^ or smaller in the discovery GWAS were included in the analysis. Variants were first ordered by ascending p-value. A variant was considered to be an independent signal if the association p-value after conditioning (conditional p-value) on the tag signal was smaller than 0.05. Newly identified independent signals were included with the tag signal for conditioning on the next variant.

### Region-based association analysis

Region-based association analyses were performed in 1 Kb sliding windows with 500 bp increments in a 1 Mb flanking region of the tag GWAS signal using the SKAT_CommonRare function from the SKAT R package v1.3.2.1 (Ionita-Laza *et al*. 2013). Default settings were used with method = “C” and test.type = “Joint”. A minor allele frequency (MAF) threshold of 0.01 was used as the cutoff to distinguish rare and common variants. Variants were annotated in TOPMed using the WGSA pipeline (Liu *et al*. 2016). Since SKAT imputes missing genotypes by default by assigning mean genotype values (impute.method=“fixed”), we chose to use low coverage genotypes instead of SKAT imputation, and hence, TOPMed freeze 8 DP0 variants with a VCF FILTER of PASS were included in the analysis. The function effectiveSize in the R package CODA (Plummer *et al*. 2006) was used to estimate the effective number of independent hypothesis tests for accurate Bonferroni multiple testing corrections. P-value thresholds for statistical significance and suggestive significance were defined as 0.05 and 1 divided by the effective number of tests, respectively (Duggal *et al*. 2008). If a region was suggestively significant, region-based analyses were repeated with functional variants and/or rare variants (MAF <= 0.01) to assess contribution of common, rare and/or functional variants. Region-based analyses using rare variants only were performed using SKAT-O (Lee *et al*. 2012). The WGSA annotation filters used to define functional variants are provided in File S1 (Supplementary Text 1). To study the contribution of individual variants to a region-based association p-value, drop-one variant analysis was performed by repeating the region-based analysis multiple times and dropping one variant only at a time.

### Functional annotations and prioritization of genetic variants

The Hi-C Unifying Genomic Interrogator (HUGIN) (Ay *et al*. 2014; Martin, J. S. *et al*. 2017; Schmitt *et al*. 2016) was used to assign potential gene targets to each variant. HUGIN uses the Hi-C data generated from the primary human tissues from four donors used in the Roadmap Epigenomics Project (Schmitt *et al*. 2016). ENCODE annotations (ENCODE Project Consortium 2011; ENCODE Project Consortium 2012) were based on overlap of the variants with functional data downloaded from the UCSC Table Browser (Karolchik *et al*. 2004). These data included DNAase I hypersensitivity peak clusters (hg38 wgEncodeRegDnaseClustered table), transcription factor ChIP-Seq clusters (hg38 encRegTfbsClustered table) and histone modification ChIP-Seq peaks (hg19 wgEncodeBroadHistone<cell type><histone>StdPk tables). For DNase I hypersensitivity and transcription factor binding sites, we focused on blood, bone marrow, lung and embryonic cells. For histone modification ChIP-Seq, we focused on H3K27ac and H3K4me3 modifications in human blood (GM12878), bone marrow (K562), lung fibroblast (NHLF), and embryonic stem cells (H1-hESC). LiftOver tool (Hinrichs *et al*. 2006) was used to convert genomic coordinates from hg19 to hg38. Candidate cis-regulatory elements (ccREs) were a subset of representative DNase hypersensitivity sites with epigenetic activity further supported by histone modification (H3K4me3 and H3K27ac) or CTCF-binding data from the ENCODE project. Overlap of variants with ccREs were detected using the Search Candidate cis-Regulatory Elements by ENCODE (SCREEN) web interface (ENCODE Project Consortium 2011; ENCODE Project Consortium 2012).

Prioritization of genetic variants was based on the presence of statistical, functional and/or bioinformatic evidence as described in the Diverse Convergent Evidence (DiCE) prioritization framework (Ciesielski *et al*. 2014). The priority score of each variant was obtained by counting the number of statistical, functional, and/or bioinformatic evidences that support potential biological function for that variant.

### Replication of GWAS associations

All replication analyses were performed in subjects with asthma. Replication of GWAS FEV_1_ associations was attempted on TOPMed whole genome sequencing data generated from four cohorts. These cohorts included Puerto Rican (n=1,109) and Mexican American (n=649) children in the Genes-Environments and Admixture in Latino Americans (GALA II) study (Oh *et al*. 2012), African American adults in the Study of Asthma Phenotypes and Pharmacogenomic Interactions by Race-Ethnicity (SAPPHIRE, n=3,428) (Levin *et al*. 2014) and African American children in Genetics of Complex Pediatric Disorders (GCPD-A, n=1,464) study (Ong *et al*. 2013). Age, sex, height, controller medications and the first 5 PCs were used as covariates.

Additionally, replication of GWAS FEV_1_ associations was attempted using data of black UK Biobank subjects who had asthma (n=627) while adjusting for age, sex, height and the first 5 principal components. Asthma status was defined by ICD code or self-reported asthma. UK Biobank genotype data was generated on Affymetrix UK BiLEVE axiom or UK Biobank Axiom array and imputed into the Haplotype Reference Consortium, 1000G and UK 10K projects (Bycroft *et al*. 2018; Canela-Xandri *et al*. 2018). Additional details on the UK Biobank study and the replication procedures are available in File S1 (Supplementary Text 2).

### RNA sequencing and expression quantitative trait loci (eQTL) analysis

Whole-transcriptome libraries of 370 nasal brushings from GALA II Puerto Rican children with asthma were constructed by using the Beckman Coulter FX automation system (Beckman Coulter, Fullerton, CA). Libraries were sequenced with the Illumina HiSeq 2500 system. Raw RNA-Seq reads were trimmed using Skewer (Jiang *et al*. 2014) and mapped to human reference genome hg38 using Hisat2 (Kim, D. *et al*. 2015). Reads mapped to genes were counted with htseq-count and using the UCSC hg38 GTF file as reference (Anders *et al*. 2015). Cis-expression quantitative trait locus (eQTL) analysis of *KITLG* was performed as described in the Genotype-Tissue Expression (GTEx) project version 7 protocol (GTEx Consortium *et al*. 2017) using age, sex, BMI, global African and European ancestries and 60 PEER factors as covariates.

### Gene-by-air-pollution interaction analysis

We hypothesized that the effect of genetic variation on lung function in our study population may differ by the levels of exposure to SO_2_ (Neophytou *et al*. 2016). To test for an interaction between a genetic variant and SO_2_, an additional multiplicative interaction term (variant × S0_2_ exposure) was included in the original GWAS model (see Method Section “FEV_1_ GWAS”). The SO_2_ estimates used in the interaction analysis were first-year, past-year, and lifetime exposure to ambient of SO_2_, which were estimated as described previously (Neophytou *et al*. 2016). Briefly, we obtained regional ambient daily air pollution data from the U.S. Environmental Protection Agency Air Quality System. SO_2_ estimates for the participant’s residential geographic coordinate were calculated as the inverse distance-squared weighted average from the four closest air pollution monitoring stations within 50 km of the participant’s residence. We estimated yearly exposure at the reported residential address by averaging all available daily measures (daily average of 1-hour SO_2_) in a given year. If the participant had a change of residential address in a given year, we estimate yearly exposure as a time-weighted estimate based on the number of months spent at each different address in that year. Average lifetime exposures were estimated using all available yearly average estimates over the lifetime of the participant until the day of spirometry testing. Since not all pollutants were measured daily, there are location-and pollutant-dependent missing values. Residuals of FEV_1_ were plotted against exposure to SO_2_ and stratified by the number of copies of the minor allele. Residuals of FEV_1_ were obtained as described in the Methods Section “FEV_1_ GWAS”.

### Data availability

Local institutional review boards approved the studies (IRB# 10-02877). All subjects and legal guardians provided written informed consent. TOPMed whole genome sequencing and phenotype data from SAGE II are available on dbGaP under accession number phs000921.v4.p1. Normalized gene count data for *KITLG* and supplemental materials are available at figshare.

## RESULTS

### Novel lung function associations

Subject characteristics of the 867 African American children with asthma included in this study are shown in Table 1, and the distribution of their FEV_1_ measurements (mean = 2.56 L, standard deviation = 0.79 L) is in Figure S1. The CODA-adjusted statistical significance thresholds 2.10 × 10^−8^ and 4.19 × 10^−7^ were used as the genome-wide and suggestive significance thresholds, respectively. According to this threshold, one SNP in chromosome 12 (chr12:88846435, rs73429450, G>A) was associated with FEV_1_.res.rnorm (Figure 1, p = 9.01 × 10^−9^, β = 0.801) at genome-wide significance. The association between rs73429450 and lung function remained statistically significant when the association was repeated using untransformed FEV_1_ (p = 1.26 × 10^−8^, β = 0.302) as the outcome variable. The association between rs73429450 and lung function was suggestive using FEV_1_.perc.predicted (p = 1.69 × 10^−7^, β = 0.100). Twenty suggestive associations corresponding to 4 tag signals are reported in Supplementary File S2. None of the suggestive associations overlapped with any of the previously reported FEV_1_-associated loci. When considering only common variants and applying a p-value threshold of 5.63 × 10^−5^, we found replicated in 6 out of 230 previously reported FEV_1_ associations (Table S1). Our top FEV_1_ association, rs73429450, did not overlap with any previously reported loci and it is a novel association with FEV_1_ in this study population.

**Table 1.**
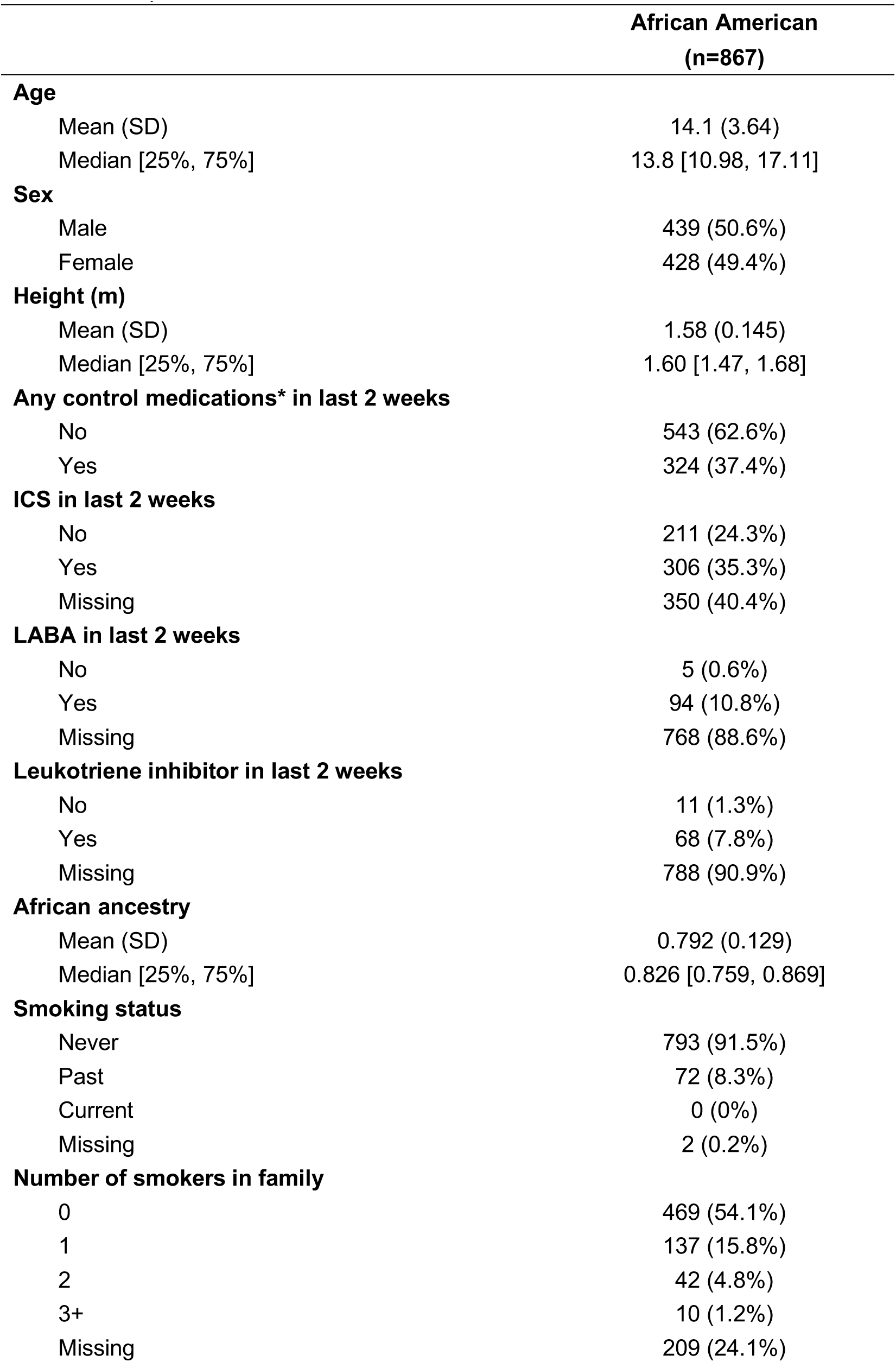
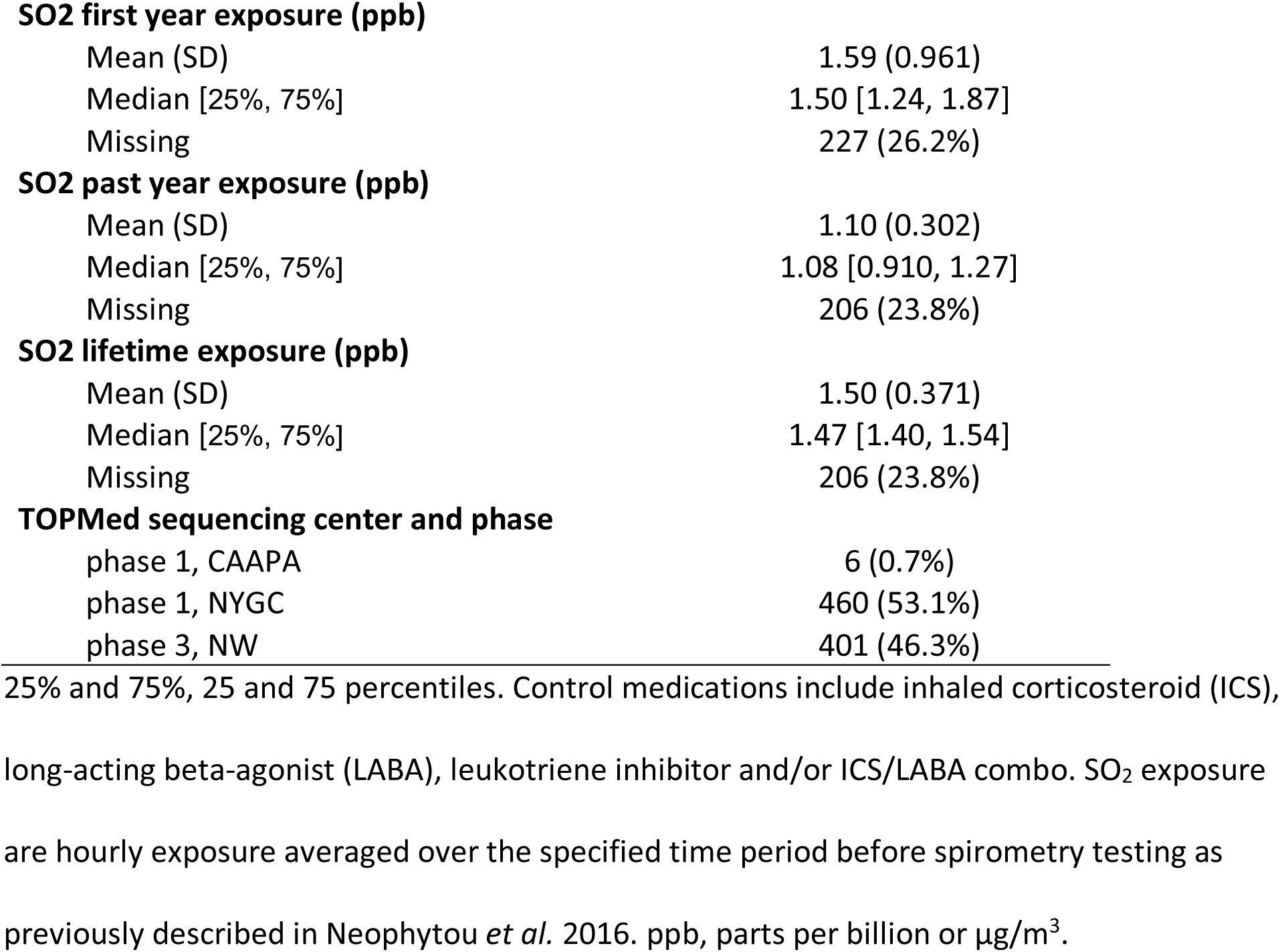
Descriptive characteristics of 867 African American children with asthma included in this study.

|  | **African American**<br>**(n=867)** |
| --- | --- |
| **Age** |  |
| Mean (SD) | 14.1 (3.64) |
| Median [25%, 75%] | 13.8 [10.98, 17.11] |
| **Sex** |  |
| Male | 439 (50.6%) |
| Female | 428 (49.4%) |
| **Height (m)** |  |
| Mean (SD) | 1.58 (0.145) |
| Median [25%, 75%] | 1.60 [1.47, 1.68] |
| **Any control medications\* in last 2 weeks** |  |
| No | 543 (62.6%) |
| Yes | 324 (37.4%) |
| **ICS in last 2 weeks** |  |
| No | 211 (24.3%) |
| Yes | 306 (35.3%) |
| Missing | 350 (40.4%) |
| **LABA in last 2 weeks** |  |
| No | 5 (0.6%) |
| Yes | 94 (10.8%) |
| Missing | 768 (88.6%) |
| **Leukotriene inhibitor in last 2 weeks** |  |
| No | 11 (1.3%) |
| Yes | 68 (7.8%) |
| Missing | 788 (90.9%) |
| **African ancestry** |  |
| Mean (SD) | 0.792 (0.129) |
| Median [25%, 75%] | 0.826 [0.759, 0.869] |
| **Smoking status** |  |
| Never | 793 (91.5%) |
| Past | 72 (8.3%) |
| Current | 0 (0%) |
| Missing | 2 (0.2%) |
| **Number of smokers in family** |  |
| 0 | 469 (54.1%) |
| 1 | 137 (15.8%) |
| 2 | 42 (4.8%) |
| 3+ | 10 (1.2%) |
| Missing | 209 (24.1%) |
| **SO2 first year exposure (ppb)** |  |
| Mean (SD) | 1.59 (0.961) |
| Median [25%, 75%] | 1.50 [1.24, 1.87] |
| Missing | 227 (26.2%) |
| **SO2 past year exposure (ppb)** |  |
| Mean (SD) | 1.10 (0.302) |
| Median [25%, 75%] | 1.08 [0.910, 1.27] |
| Missing | 206 (23.8%) |
| **SO2 lifetime exposure (ppb)** |  |
| Mean (SD) | 1.50 (0.371) |
| Median [25%, 75%] | 1.47 [1.40, 1.54] |
| Missing | 206 (23.8%) |
| **TOPMed sequencing center and phase** |  |
| phase 1, CAAPA | 6 (0.7%) |
| phase 1, NYGC | 460 (53.1%) |
| phase 3, NW | 401 (46.3%) |

25% and 75%, 25 and 75 percentiles. Control medications include inhaled corticosteroid (ICS), long-acting beta-agonist (LABA), leukotriene inhibitor and/or ICS/LABA combo. SO<sub>2</sub> exposure are hourly exposure averaged over the specified time period before spirometry testing as previously described in Neophytou *et al.* 2016. ppb, parts per billion or µg/m<sup>3</sup>.

**Figure 1.**
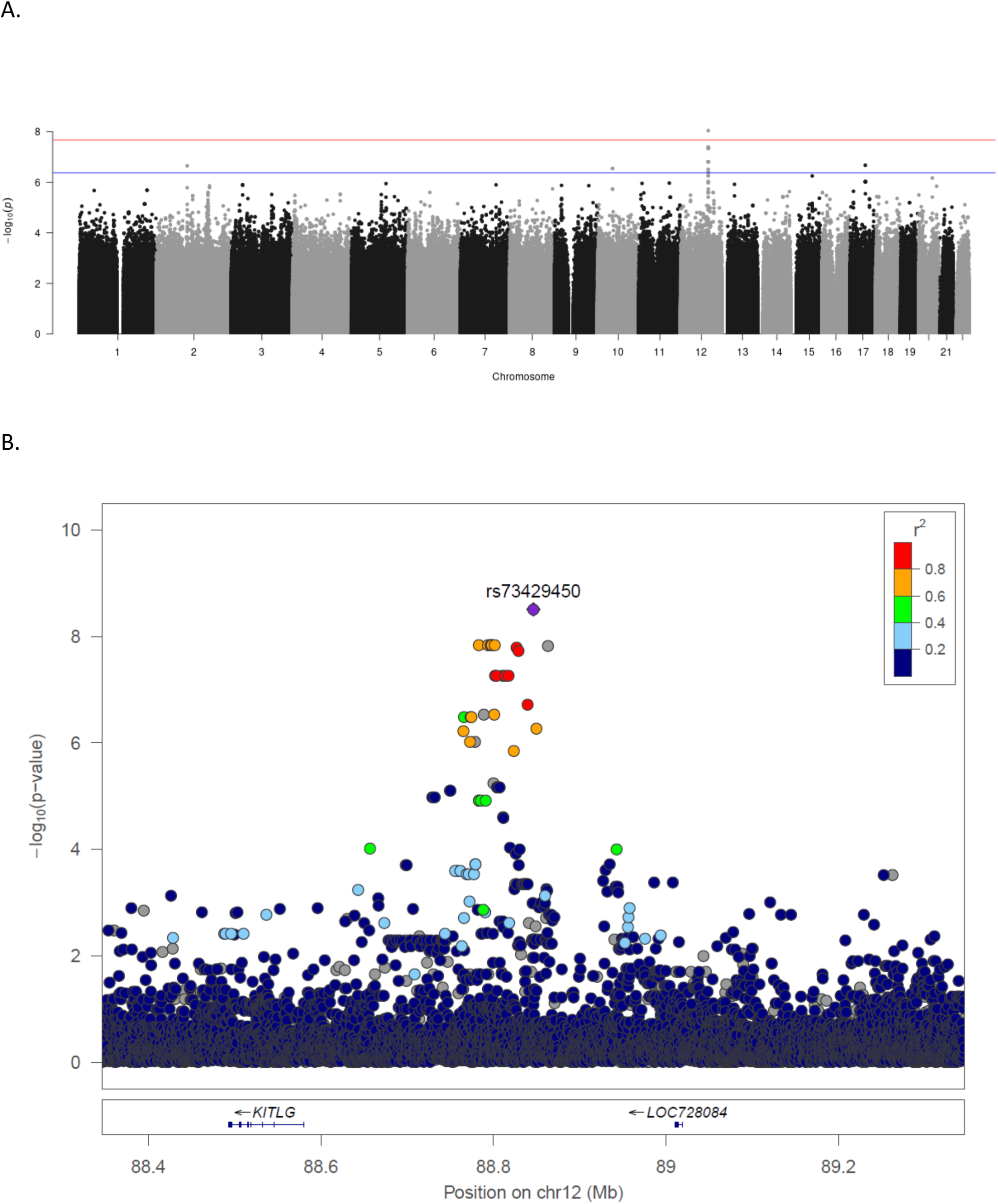
Manhattan and LocusZoom plots from genome-wide association study of lung function*. (A) Manhattan plot from genome-wide association study of lung function* using linear regression in ENCORE. Red horizontal line: CODA-adjusted genome-wide significance p-value of 2.10 × 10^−8^. Blue horizontal line: CODA-adjusted suggestive significance p-value of 4.19 × 10^−7^. (B) LocusZoom plot of rs73429450 (chr12 : 88846435) and 500 Kb flanking region. Colors show linkage disequilibrium in the study population. * FEV_1_.res.rnorm was used as the phenotype for the association testing.

Secondary analysis that included covariates correcting for smoking status and number of smokers in the family showed that smoking-related factors were not significantly associated with FEV_1_ in our pediatric SAGE cohort: using 657 out of 867 individuals with available smoking-related covariates, the FEV_1_.res.rnorm association p-values before and after including the smoking-related covariates were 2.01 × 10^−6^ and 1.89 × 10^−6^. Both p-values of the covariates smoking status (p = 0.27) and number of smokers in the family (p = 0.54) were not significant.

Conditional analysis was performed on 45 variants with association p < 1 × 10^−4^located within 1 Mb of the strongest association signal (rs73429450). Two weaker independent signals (rs17016065, rs58475486) were identified (Table S2). None of the 45 variants showed association with FEV_1_.res.rnom in 251 SAGE II children without asthma (Table S3).

The minor allele frequency of rs73429450 in continental populations from the 1000 Genomes Project (1000G) is 3% in Africans (AFR) and < 1% in Admixed Americans (AMR), Europeans (EUR) and Asians (EAS and SAS) (1000 Genomes Project Consortium *et al*. 2015). Rs73429450 was not included on the Affymetrix LAT1 genotyping array where SAGE participants were previously genotyped. To determine if the rs73429450 association with FEV_1_ was only identifiable using whole genome sequencing data, we attempted to reproduce our results by imputing the genotype of rs73429450 in 851 SAGE participants with available array data using 1000G phase 3 (n = 2,504), HRC r1.1 (n = 32,470), CAAPA (n = 883) and TOPMed freeze 5 (n = 62,784) reference panels. Our results remained statistically significant when using the 1000G phase 3 (p = 4.97 × 10^−8^, β = 0.79, imputation R^2^ = 0.95) and TOPMed freeze 5 (p = 1.22 × 10^−8^, β = 0.80, imputation R^2^ = 0.98) reference panels, but lost statistical significance when rs73429450 genotypes were imputed using the HRC (p = 4.35 × 10^−7^, β = 0.68, imputation R^2^ = 0.94) and CAAPA (p = 1.95 × 10^−7^, β = 0.80, imputation R^2^ = 0.71) reference panels.

Region-based association analysis including all variants conditioned on the association signal from rs73429450 was performed in its 1 Mb flanking region (chr12:87846435-89846435). No windows were significantly associated after Bonferroni multiple testing correction (p < 2.80 × 10^−4^, Figure S2), but 20 windows were suggestively associated with FEV_1_.res.rnorm (p < 5.60 × 10^−3^, Table S5). Two of 20 windows re-tested using only functional variants were suggestively significant (region 4 and 16). Both of these windows were no longer suggestively significant after removing the common variants, indicating that association signal from these regions was mostly driven by common variants. Further investigation on region 16 using drop-one analysis on the 2 rare and 1 common function variants confirmed the major contribution by the common variant, rs1895710, as shown by the major increase in p-value (Table S6). The signal was also slightly driven by the singleton, rs990979778. Drop-one analysis was not performed on region 4 because there were only 1 common and 1 rare variants.

A Hi-C assay couples a chromosome conformation capture (3C) assay with next-generation sequencing to capture long-range interactions in the genome. We identified a statistically significant long-range chromatin interaction between the GWAS peak and the KIT ligand (*KITLG*, also known as stem cell factor, *SCF*) gene in human fetal lung fibroblast cell line IMR90 (Table S7). The long-range interaction detected in human primary lung tissue was not significant, implying that the potential long-range interactions are specific to tissue type or developmental stage.

### Potential regulatory role of FEV_1_-associated variants on KITLG expression

To further elucidate potential regulatory relationships between the GWAS association peak and *KITLG*, we analyzed whether variants in the peak were eQTL of *KITLG* in previously published whole blood RNA-Seq data available from the same study participants (Mak, Angel CY *et al*. 2016). The whole blood RNA-Seq data, however, did not yield evidence of expressed *KITLG*, consistent with results in GTEx. We subsequently used RNA-Seq data from nasal epithelial cells of 370 Puerto Rican children with asthma from the GALA II study, and found that five out of 45 variants were eQTL of *KITLG* (Table S8). While Puerto Ricans are a different population than African Americans, they are both admixed populations with substantial African genetic ancestry, and therefore could share eQTLs. All five eQTLs corresponded to one signal in a region with strong linkage disequilibrium (r^2^ > 0.8, Figure S3).

### Replication of genetic association with FEV_1_

Subject characteristics of our four replication cohorts (SAPPHIRE, GCPD-A, UK Biobank and GALA II) are shown in Table S9. We attempted to replicate the association of the 45 SNPs in our primary FEV_1_ GWAS in each cohort. We used 0.05 as the suggestive p-value threshold and 0.0167 as the Bonferroni-corrected p-value threshold after correcting for 3 independent signals (see conditional analysis in Results Section). A total of 20 variants were replicated at p < 0.05 with consistent direction of effect in black UK Biobank participants; 14 variants in SAPPHIRE and 2 variants in GCPD-A were significant but had an opposite direction of effect (Table S10).

We attempted to replicate the FEV_1_.res.rnorm association in Mexican American (n = 649) and Puerto Rican (n = 1,109) children with asthma from the GALA II study. In Mexican Americans, we excluded 19 variants with MAF < 0.1% and associations for the remaining 26 variants did not replicate (Table S11). In Puerto Ricans, the associations were not replicated (Table S11).

### Incorporating statistical and functional evidence for candidate variant prioritization

We combined and summarized all functional evidence for the top 45 variants, along with eQTL findings from nasal epithelial RNA-Seq and replication results (Figure 2, Table 2 and S12). To facilitate interpretation of the variant association with FEV_1_, the effect sizes and p-values of both FEV_1_ (β and p) and FEV_1_.res.rnorm (β_norm_ and p_norm_) associations are also reported. CADD functional prediction score and ENCODE histone modification ChIP-Seq peaks in embryonic, blood, bone marrow, and lung-related tissues were also examined but not reported because none of the variants had a CADD score greater than 10 and none overlapped with histone modification sites. Rs73440122 received the highest priority score of 3 based on replication in the UK Biobank, overlap with a DNase I hypersensitivity site in B-lymphoblastoid cells (GM12865) and overlap with an SPI1 binding site in acute promyelocytic leukemia cells. Eight other variants were prioritized with score > 2 or evidence of being an eQTL for *KITLG* in nasal epithelial cells (Table 2, score marked with ^ or # respectively). These nine candidate variants were selected for gene-by-air-pollution interaction analyses.

**Table 2.** Genome-wide lung function association in SAGE II children with asthma.

| M | rsID | Alt | $\beta$ | p | $\beta_{\text{norm}}$ | $p_{\text{norm}}$ | MAF | 1000 Genomes | | | | Score |
| --- | --- | --- | --- | --- | --- | --- | --- | --- | --- | --- | --- | --- |
|  |  |  |  |  |  |  |  | ALL | AFR | AMR | EUR |  |
| 1 | rs11835305 | T | 0.126 | 1.93E-05 | 0.320 | 3.69E-05 | 0.104 | 0.036 | 0.119 | 0.012 | 0.001 | 0 |
| 2 | rs17015963 | C | 0.126 | 1.93E-05 | 0.320 | 3.69E-05 | 0.104 | 0.036 | 0.120 | 0.012 | 0.001 | 0 |
| 3 | rs58475486* | T | 0.127 | 1.45E-05 | 0.323 | 2.81E-05 | 0.105 | 0.037 | 0.123 | 0.012 | 0.001 | 2^ |
| 4 | rs17015979 | T | 0.127 | 1.45E-05 | 0.323 | 2.81E-05 | 0.105 | 0.037 | 0.123 | 0.012 | 0.001 | 0 |
| 5 | rs57692452 | C | 0.245 | 1.63E-06 | 0.654 | 1.06E-06 | 0.033 | 0.011 | 0.030 | 0.006 | 0.001 | 2^ |
| 6 | rs112585732 | T | 0.235 | 4.35E-07 | 0.625 | 3.06E-07 | 0.041 | 0.016 | 0.050 | 0.006 | 0.001 | 0 |
| 7 | rs113837356 | T | 0.270 | 3.44E-06 | 0.719 | 2.49E-06 | 0.025 | 0.008 | 0.027 | 0.006 | 0.001 | 1 |
| 8 | rs61441836 | G | 0.252 | 8.19E-07 | 0.671 | 5.46E-07 | 0.033 | 0.010 | 0.030 | 0.006 | 0.001 | 1 |
| 9 | rs73438172 | A | 0.252 | 8.19E-07 | 0.671 | 5.46E-07 | 0.033 | 0.010 | 0.030 | 0.006 | 0.001 | 1 |
| 10 | rs1044043958& | A | 0.270 | 3.44E-06 | 0.719 | 2.49E-06 | 0.025 | - | - | - | - | 0 |
| 11 | rs73438182 | G | 0.138 | 1.68E-04 | 0.378 | 9.18E-05 | 0.064 | 0.020 | 0.068 | 0.006 | 0.001 | 0 |
| 12 | rs73438185 | A | 0.138 | 1.68E-04 | 0.378 | 9.18E-05 | 0.064 | 0.020 | 0.068 | 0.006 | 0.001 | 0 |
| 13 | rs73438188 | A | 0.297 | 6.97E-08 | 0.792 | 4.42E-08 | 0.028 | 0.010 | 0.027 | 0.006 | 0.001 | 1 |
| 14 | rs73438190 | C | 0.181 | 1.86E-05 | 0.486 | 1.22E-05 | 0.048 | 0.016 | 0.047 | 0.006 | 0.001 | 0 |
| 15 | rs73438195 | A | 0.181 | 1.86E-05 | 0.486 | 1.22E-05 | 0.048 | 0.016 | 0.047 | 0.006 | 0.001 | 0 |
| 16 | rs111857459 | T | 0.181 | 1.86E-05 | 0.486 | 1.22E-05 | 0.048 | 0.016 | 0.047 | 0.006 | 0.001 | 0 |
| 17 | rs144369986& | T | 0.285 | 1.21E-06 | 0.756 | 9.44E-07 | 0.025 | 0.008 | 0.026 | 0.006 | 0.001 | 0 |
| 18 | rs73440106 | G | 0.181 | 1.86E-05 | 0.486 | 1.22E-05 | 0.048 | 0.016 | 0.047 | 0.006 | 0.001 | 0 |
| 19 | rs73440107 | A | 0.297 | 6.97E-08 | 0.792 | 4.42E-08 | 0.028 | 0.010 | 0.027 | 0.006 | 0.001 | 1 |
| 20 | rs111453514 | C | 0.297 | 6.97E-08 | 0.792 | 4.42E-08 | 0.028 | 0.010 | 0.027 | 0.006 | 0.001 | 1 |
| 21 | rs73440112 | T | 0.297 | 6.97E-08 | 0.792 | 4.42E-08 | 0.028 | 0.010 | 0.027 | 0.006 | 0.001 | 1 |
| 22 | rs73440115 | G | 0.297 | 6.97E-08 | 0.792 | 4.42E-08 | 0.028 | 0.011 | 0.028 | 0.006 | 0.001 | 1 |
| 23 | rs11312747& | A | 0.133 | 1.43E-05 | 0.357 | 8.51E-06 | 0.100 | 0.036 | 0.121 | 0.010 | 0.001 | 0 |
| 24 | rs73440120 | A | 0.285 | 1.21E-06 | 0.756 | 9.44E-07 | 0.025 | 0.008 | 0.026 | 0.006 | 0.001 | 1 |
| 25 | rs111289668 | G | 0.297 | 6.97E-08 | 0.792 | 4.42E-08 | 0.028 | 0.010 | 0.027 | 0.006 | 0.001 | 2^ |
| 26 | rs73440122 | C | 0.292 | 2.08E-07 | 0.775 | 1.55E-07 | 0.027 | 0.011 | 0.030 | 0.006 | 0.001 | 3^ |
| 27 | rs73440123 | G | 0.292 | 2.08E-07 | 0.775 | 1.55E-07 | 0.027 | 0.011 | 0.030 | 0.006 | 0.001 | 1 |
| 28 | rs17016065* | G | 0.112 | 3.19E-06 | 0.296 | 2.54E-06 | 0.177 | 0.075 | 0.217 | 0.017 | 0.009 | 1# |
| 29 | rs17016066 | A | 0.112 | 3.19E-06 | 0.296 | 2.54E-06 | 0.177 | 0.075 | 0.217 | 0.017 | 0.009 | 1# |
| 30 | rs147400083& | T | 0.112 | 3.19E-06 | 0.296 | 2.54E-06 | 0.177 | - | - | - | - | 0 |
| 31 | rs866852270 | T | 0.112 | 3.19E-06 | 0.296 | 2.54E-06 | 0.177 | - | - | - | - | 0 |
| 32 | rs141293300& | C | 0.292 | 2.08E-07 | 0.775 | 1.55E-07 | 0.027 | 0.011 | 0.030 | 0.006 | 0.001 | 1 |
| 33 | rs1398303 | A | 0.104 | 1.22E-05 | 0.274 | 1.12E-05 | 0.186 | 0.077 | 0.223 | 0.020 | 0.009 | 1# |
| 34 | rs61924868 | T | 0.104 | 1.24E-05 | 0.275 | 1.14E-05 | 0.185 | 0.078 | 0.223 | 0.020 | 0.009 | 1# |
| 35 | rs73440134 | T | 0.292 | 2.08E-07 | 0.775 | 1.55E-07 | 0.027 | 0.011 | 0.030 | 0.006 | 0.001 | 1 |
| 36 | rs73429413 | G | 0.292 | 2.08E-07 | 0.775 | 1.55E-07 | 0.027 | 0.011 | 0.030 | 0.006 | 0.001 | 1 |
| 37 | rs73429415 | A | 0.096 | 5.13E-05 | 0.253 | 4.84E-05 | 0.189 | 0.078 | 0.225 | 0.022 | 0.009 | 1# |
| 38 | rs112449284 | T | 0.242 | 4.64E-06 | 0.640 | 3.87E-06 | 0.031 | 0.012 | 0.035 | 0.007 | 0.001 | 1 |
| 39 | rs111981782 | C | 0.296 | 5.78E-08 | 0.786 | 4.09E-08 | 0.029 | 0.012 | 0.033 | 0.006 | 0.001 | 1 |
| 40 | rs150942400 | T | 0.293 | 6.01E-08 | 0.780 | 4.01E-08 | 0.029 | 0.012 | 0.034 | 0.006 | 0.002 | 1 |
| 41 | rs147527487 | C | 0.086 | 8.49E-05 | 0.226 | 8.14E-05 | 0.205 | 0.095 | 0.249 | 0.016 | 0.005 | 0 |
| 42 | rs111243672 | A | 0.258 | 6.41E-07 | 0.690 | 3.99E-07 | 0.032 | 0.014 | 0.037 | 0.007 | 0.004 | 1 |
| 43 | rs73429450* | A | 0.302 | 1.26E-08 | 0.801 | 9.01E-09 | 0.031 | 0.012 | 0.033 | 0.009 | 0.002 | 1 |
| 44 | rs758775577 | C | 0.217 | 2.22E-06 | 0.574 | 1.85E-06 | 0.041 | - | - | - | - | 0 |
| 45 | rs142679473<sup>&</sup> | C | 0.285 | 6.30E-08 | 0.756 | 4.62E-08 | 0.031 | 0.012 | 0.033 | 0.009 | 0.002 | 0 |

Score, priority score based on statistical and functional evidences which are reported in Table

**Figure 2.**
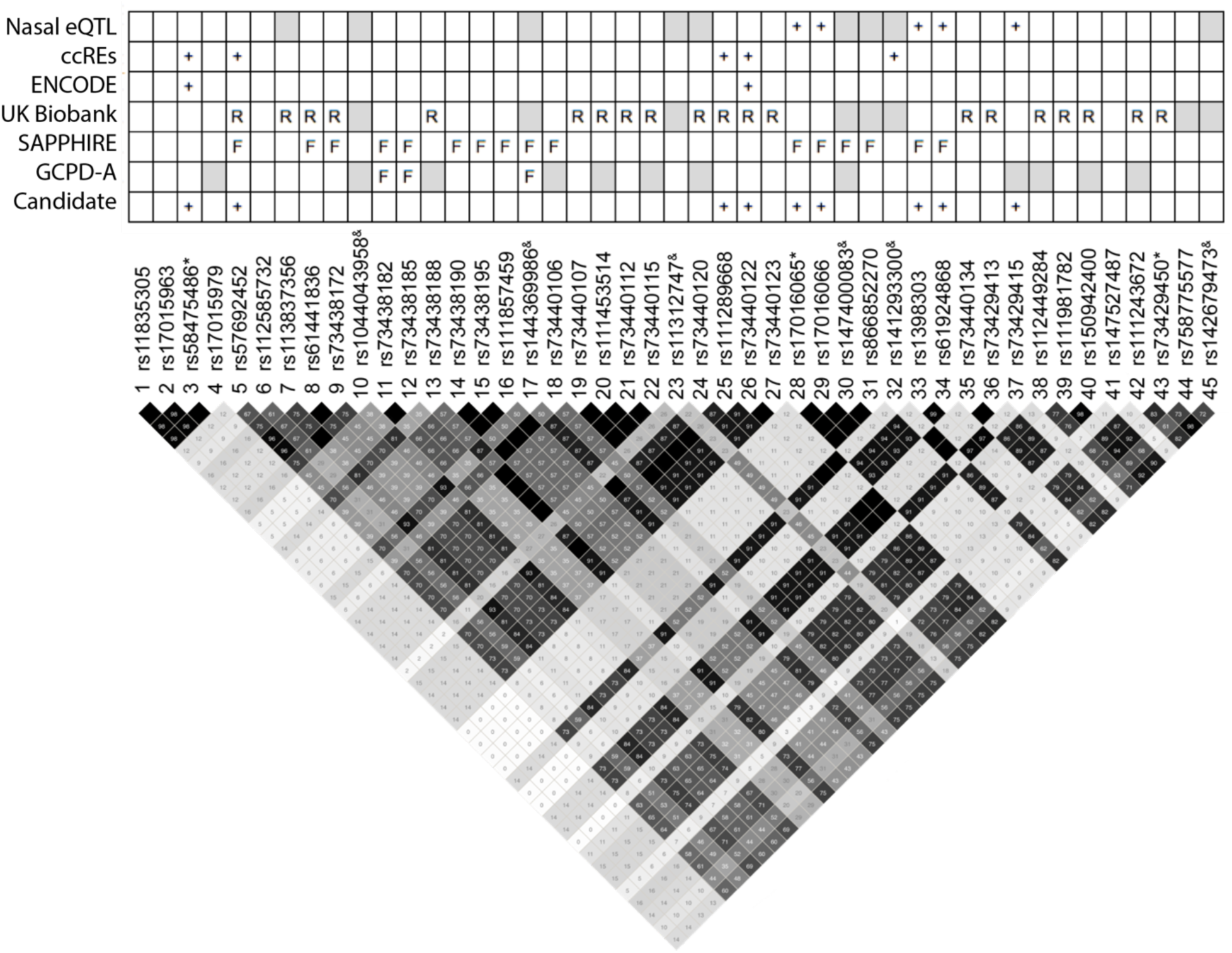
Integration of statistical and functional evidence for variant prioritization. Numbers and different shades of black in the LD plot represent LD in R^2^. The three independent signals identified in the conditional analysis are marked with *. Indels are marked with ^&^. Nasal eQTL, variants eQTL of *KITLG* in nasal epithelial cells. ccREs, candidate cis-regulatory elements in SCREEN registry. ENCORE, DNase I hypersensitivity site and/or transcription factor ChIP-Seq overlapping with the variants. UK Biobank, SAPPHIRE, GCPD-A, replication results using Blacks in UK Biobank and African Americans in the SAPPHIRE and GCPD-A cohorts (R = replicated at p<0.05; F = flip-flop association at p < 0.05). Candidate, candidate variants prioritized because of presence of two or more evidence or is nasal eQTL. + indicates presence of evidence. Boxes in the top panel were shaded grey if results were not available.

### Gene-by-air-pollution interaction of rs58475486

We previously found that first year of life and lifetime exposure to SO_2_ were associated with FEV_1_ in African American children (Neophytou *et al*. 2016). We investigated whether the effect of the nine prioritized genetic variants associated with lung function varied by SO_2_ exposure (first year of life, past year, and lifetime exposure). Since the nine variants represent three independent signals (see conditional analysis in the Results Section), the Bonferroni-corrected p-value threshold was set to p = 0.0056 (correction for nine tests; three signals and three exposure periods to SO_2_). We observed a single statistically significant interaction between the T allele of rs58475486 and past year exposure to SO_2_ that was positively associated with FEV_1_ (p = 0.003, β = 0.32, Table 3, Figure 3A). This interaction remains significant (p = 0.003, β = 0.32) in secondary analyses adjusted for smoking status or a multiplicative interaction term of rs58475486 and smoking status as additional covariates. Interestingly, six of the remaining eight variants also displayed interaction effects with past year exposure to SO_2_ that were suggestively associated (p< 0.05) with FEV_1_ (Table 3). We also found a suggestive interaction of the C allele of rs73440122 with first year exposure to SO_2_ that was associated with decreased FEV_1_ (p = 0.045, β = −0.32, Figure 3B). The same allele also showed interaction with past year of exposure to SO_2_ that was suggestively associated with FEV_1_ in the opposite direction (p = 0.051, β = 0.39).

**Table 3.** Gene-and-environment analysis on FEV_1_

| Variant | Exposure | n | Variant |  | Exposure |  | GxE |  |
| --- | --- | --- | --- | --- | --- | --- | --- | --- |
| | | | $\beta$ | p | $\beta$ | p | $\beta$ | p |
| rs58475486_T<sup>1</sup> | SO<sub>2</sub> first year | 640 | 0.13 | 6.62E-06 | -0.05 | 0.003 | -0.03 | 0.658 |
| rs57692452_C<sup>2</sup> | SO<sub>2</sub> first year | 640 | 0.25 | 1.16E-06 | -0.05 | 0.003 | -0.24 | 0.091 |
| rs111289668_G<sup>2</sup> | SO<sub>2</sub> first year | 640 | 0.31 | 2.53E-08 | -0.05 | 0.003 | -0.27 | 0.079 |
| rs73440122_C<sup>2</sup> | SO<sub>2</sub> first year | 640 | 0.31 | 7.78E-08 | -0.05 | 0.003 | -0.32 | 0.045 * |
| rs17016065_G<sup>3</sup> | SO<sub>2</sub> first year | 640 | 0.11 | 8.82E-06 | -0.05 | 0.003 | -0.08 | 0.108 |
| rs17016066_A<sup>3</sup> | SO<sub>2</sub> first year | 640 | 0.11 | 8.82E-06 | -0.05 | 0.003 | -0.08 | 0.108 |
| rs1398303_A<sup>3</sup> | SO<sub>2</sub> first year | 640 | 0.10 | 3.15E-05 | -0.05 | 0.003 | -0.09 | 0.082 |
| rs61924868_T<sup>3</sup> | SO<sub>2</sub> first year | 640 | 0.10 | 3.30E-05 | -0.05 | 0.003 | -0.08 | 0.088 |
| rs73429415_A<sup>3</sup> | SO<sub>2</sub> first year | 640 | 0.09 | 1.09E-04 | -0.05 | 0.003 | -0.09 | 0.069 |
| rs58475486_T<sup>1</sup> | SO<sub>2</sub> past year | 661 | 0.13 | 6.62E-06 | 0.05 | 0.362 | 0.32 | 0.003 ** |
| rs57692452_C<sup>2</sup> | SO<sub>2</sub> past year | 661 | 0.25 | 1.16E-06 | 0.05 | 0.362 | 0.29 | 0.100 |
| rs111289668_G<sup>2</sup> | SO<sub>2</sub> past year | 661 | 0.31 | 2.53E-08 | 0.05 | 0.362 | 0.41 | 0.037 * |
| rs73440122_C<sup>2</sup> | SO<sub>2</sub> past year | 661 | 0.31 | 7.78E-08 | 0.05 | 0.362 | 0.39 | 0.051 |
| rs17016065_G<sup>3</sup> | SO<sub>2</sub> past year | 661 | 0.11 | 8.82E-06 | 0.05 | 0.362 | 0.20 | 0.026 * |
| rs17016066_A<sup>3</sup> | SO<sub>2</sub> past year | 661 | 0.11 | 8.82E-06 | 0.05 | 0.362 | 0.20 | 0.026 * |
| rs1398303_A<sup>3</sup> | SO<sub>2</sub> past year | 661 | 0.10 | 3.15E-05 | 0.05 | 0.362 | 0.21 | 0.021 * |
| rs61924868_T<sup>3</sup> | SO<sub>2</sub> past year | 661 | 0.10 | 3.30E-05 | 0.05 | 0.362 | 0.21 | 0.023 * |
| rs73429415_A<sup>3</sup> | SO<sub>2</sub> past year | 661 | 0.09 | 1.09E-04 | 0.05 | 0.362 | 0.20 | 0.026 * |
| rs58475486_T<sup>1</sup> | SO<sub>2</sub> lifetime | 661 | 0.13 | 6.62E-06 | -0.13 | 0.001 | 0.26 | 0.173 |
| rs57692452_C<sup>2</sup> | SO<sub>2</sub> lifetime | 661 | 0.25 | 1.16E-06 | -0.13 | 0.001 | 0.47 | 0.221 |
| rs111289668_G<sup>2</sup> | SO<sub>2</sub> lifetime | 661 | 0.31 | 2.53E-08 | -0.13 | 0.001 | 0.32 | 0.444 |
| rs73440122_C<sup>2</sup> | SO<sub>2</sub> lifetime | 661 | 0.31 | 7.78E-08 | -0.13 | 0.001 | 0.29 | 0.489 |
| rs17016065_G<sup>3</sup> | SO<sub>2</sub> lifetime | 661 | 0.11 | 8.82E-06 | -0.13 | 0.001 | -0.19 | 0.143 |
| rs17016066_A<sup>3</sup> | SO<sub>2</sub> lifetime | 661 | 0.11 | 8.82E-06 | -0.13 | 0.001 | -0.19 | 0.143 |
| rs1398303_A<sup>3</sup> | SO<sub>2</sub> lifetime | 661 | 0.10 | 3.15E-05 | -0.13 | 0.001 | -0.17 | 0.184 |
| rs61924868_T<sup>3</sup> | SO<sub>2</sub> lifetime | 661 | 0.10 | 3.30E-05 | -0.13 | 0.001 | -0.17 | 0.199 |
| rs73429415_A<sup>3</sup> | SO<sub>2</sub> lifetime | 661 | 0.09 | 1.09E-04 | -0.13 | 0.001 | -0.16 | 0.207 |

\*, p < 0.05; \*\*, p < bonferroni p value of 0.0056 for GxE analysis. n, sample sizes for the gene-by-SO<sub>2</sub> interaction analysis. Superscript 1 to 3 in the variant column, variants that are in LD with the 3 independent signals, rs58475486, rs73429450 and rs17016065, respectively. $\beta$ , effect sizes from the main effects of the variants, exposure and GxE interaction, respectively.

**Figure 3.**
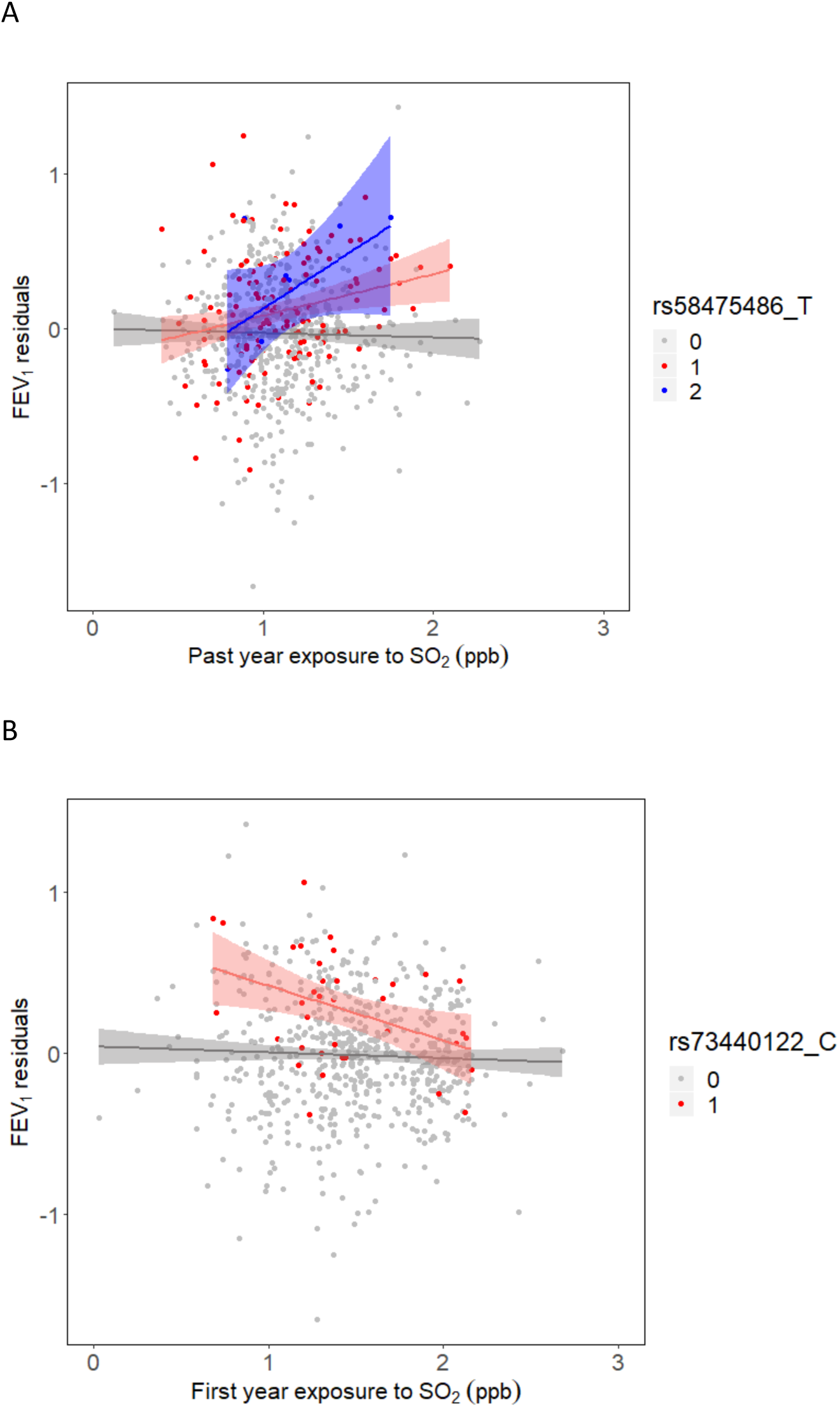
Gene-by-environment interaction analysis on FEV_1_. FEV_1_ residuals, residuals after FEV_1_ was regressed on the covariates age, sex, height, controller medications, sequencing centers and the first 5 genetic PCs. FEV_1_ residuals was plotted against (A) past year exposure to SO_2_ stratified by the number of copies of T allele of rs58475486, (B) first year of life exposure to SO_2_ stratified by the number of copies of C allele of rs73440122.

## DISCUSSION

Variant rs73429450 (MAF = 0.030) was identified as the strongest association signal with FEV_1_. Each additional copy of the protective A allele of rs73429450 was associated with a 0.3 L increase of FEV_1_. We did not find any statistically significant contribution of rare variants to the association signal from a 1 Kb sliding window analyses in the 1 MB flanking region centered on rs73429450. We were surprised to identify a novel common variant (MAF = 0.030) associated with lung function using whole genome sequence data in a population that was previously analyzed for associations with lung function using genotype array data. Further investigation revealed that our discovered variant, rs73429450, was not captured by the LAT 1 genotyping array, and the association with lung function depended on the reference panel used to impute the variant into our population. More surprisingly, our statistically significant finding was only found to be suggestively significant using data imputed from the CAAPA reference panel (p = 1.95 × 10^−7^, β = 0.80). Of the imputation reference panels that we assessed, CAAPA is one of the more relevant reference panels for our study population because it is based on African populations in the Americas. However, we note that the effect size estimated from CAAPA-imputed data was comparable to that generated from WGS data. While whole genome sequencing data is usually praised for enabling analysis of rare-variant contributions to phenotype variability, our results show the utility of whole genome sequencing data for the reliable analysis of common variants as well in the absence of relevant imputation panels.

Although rs73429450 had the lowest p-value from our whole genome sequencing association analysis, we did not find the required amount of functional evidence to prioritize this marker for inclusion in downstream gene-by-air-pollution analyses. Another variant, rs73440122, was in moderate to strong linkage disequilibrium (r^2^ = 0.76) with rs7349450 and had a similar MAF (0.027) in our study population, but was only suggestively associated with FEV_1_ in our association analysis (p = 2.08 × 10^−7^, Table2). In contrast to rs73429450, there were multiple lines of evidence suggesting the functional relevance of rs73440122: rs73440122 received the highest priority score based on its replicated FEV_1_ association in black UK Biobank participants and overlap with ENCODE gene regulatory regions, making it one of the most likely drivers of FEV_1_ variability among individuals, possibly mediated through *KITLG*.

Bioinformatic interrogation of rs73440122 revealed that the variant overlapped with a ccRE (SCREEN accession EH37E0279310), DNase I hypersensitivity site, and SPI1 ChIP-Seq clusters that were indicative of a candidate open chromatin gene regulatory region (Table S12). The binding evidence of SPI1 is highly relevant to the role of KITLG in type 2 inflammation (see below). Variant rs73440122 is located in a region that physically interacted with *KITLG* based on Hi-C data in fetal lung fibroblast cells. Additionally, five neighboring FEV_1_ associated variants were identified as eQTLs of *KITLG*, although they appeared to be an independent signal (r^2^ < 0.2). Overall, these results support regulatory interactions between our novel locus and *KITLG*.

Atopic or type 2 high asthma is the most common form of asthma in children (Comberiati *et al*. 2017). *KITLG*, more commonly known as stem cell factor (*SCF*), is a ligand of the KIT tyrosine kinase receptor. It plays an important role in type 2 inflammation in atopic asthma, especially in inflammatory processes mediated through mast cells, IgE and group 2 innate lymphoid cells (Da Silva and Frossard. 2005; Da Silva *et al*. 2006; Fonseca *et al*. 2019; Oliveira and Lukacs. 2003). In the airways, *KITLG* is expressed in bronchial epithelial cells, lung fibroblasts, bronchial smooth muscle cells, endothelial cells, peripheral blood eosinophils, dendritic cells and mast cells (Hsieh *et al*. 2005; Kassel *et al*. 1999; Oriss *et al*. 2014; Valent *et al*. 1992; Wen *et al*. 1996). *KITLG* is a major growth factor of mast cells (Reviewed in Broudy 1997; Da Silva *et al*. 2006; Galli *et al*. 1994; Galli *et al*. 1995). It promotes recruitment of mast cell progenitors into tissues (Reviewed in Oliveira and Lukacs. 2003), prevents mast cell apoptosis (Iemura *et al*. 1994; Mekori *et al*. 1993) and promotes release of inflammatory mediators such as proteases, histamine, chemotactic factors, cytokines (Reviewed in Amin 2012; Borish and Joseph. 1992). While KITLG promotes the production of cytokines like IL-13 upon IgE-receptor crosslinking on the surface of mast cells (Kobayashi *et al*. 1998), IL-13 was also reported to up-regulate *KITLG* (Rochman *et al*. 2015). Consistent with the critical role of KITLG for mast cells and type 2 inflammation, we found our prioritized variant, rs73440122, overlapped with a SPI1 (aka PU.1) ChIP-Seq cluster. The transcription factor SPI1 was demonstrated in SPI1 knockout mice to be necessary for the development of B cells, T cells, neutrophils, macrophages, dendritic cells, and mast cells (Anderson *et al*. 2000; Guerriero *et al*. 2000; McKercher *et al*. 1996; Scott *et al*. 1994; Scott *et al*. 1997; Walsh *et al*. 2002). It plays an essential role in macrophage differentiation in asthmatic and other allergic inflammation (Qian *et al*. 2015; Yashiro *et al*. 2019). It was also shown to regulate the cell fate between mast cells and monocytes (Ito *et al*. 2005; Ito *et al*. 2009; Nishiyama, Nishiyama, Ito, Masaki, Maeda *et al*. 2004; Nishiyama, Nishiyama, Ito, Masaki, Masuoka *et al*. 2004). The presence of a SPI1 binding site in a candidate regulatory region of KITLG is therefore highly relevant given the critical role of KITLG in mast cell survival and activation.

Higher levels of KITLG (Al-Muhsen *et al*. 2004; Da Silva *et al*. 2006; Tayel *et al*. 2017) and an increased number of mast cells in the lung (Cruse and Bradding. 2016; Fajt and Wenzel. 2013; Mendez-Enriquez and Hallgren. 2019) were detected in individuals with asthma. The percentage of a subpopulation of circulating blood mast cell progenitors (Lin^+^ CD34^hi^ CD117^int/hi^ FcεRI^+^) was higher in individuals with a reduced lung function (Dahlin *et al*. 2016). These findings suggested that higher *KITLG* expression and/or number of mast cells may be a contributing factor to lower lung function. This notion was inconsistent with the association of our novel locus with higher *KITLG* expression and increased lung function in SAGE II children with asthma. Interestingly, a study of 20 subjects with severe asthma found that increased in the number of chymase-positive mast cells in the small airway was associated with increased in lung function (Balzar *et al*. 2005). Overall, while there is still controversy on the direction of effect, previous findings support the association of our novel *KITLG* locus with lung function, especially in patients with allergic asthma. Our novel locus likely represents part of a complex regulatory mechanism that modulates immune cell differentiation, survival, and activation in highly cell-specific and context-dependent manners. Further studies are required to study how this locus is regulated in different airway and immune cells to affect lung function outcome in the context of asthma.

GxE interactions likely account for a portion of the “missing” heritability of many complex phenotypes (Moore and Williams. 2009). We previously found that lung function in SAGE II participants was associated with first year of life and lifetime exposures to SO_2_ (1.66% decrease [95% CI = −2.92 to −0.37] for first year of life and 5.30% decrease [95% CI = −8.43 to −2.06] for lifetime exposures in FEV_1_ per 1 ppb increases in SO_2_) (Neophytou *et al*. 2016). We hypothesized that a significant portion of the heritability of lung function was due in part to gene-by-air-pollution (SO_2_) interaction effects. The interaction between rs58475486 and past year exposure to SO_2_ that was significantly associated with lung function supports our hypothesis. The T allele of rs58475486 is common (8-14%) in African populations and showed a protective effect on lung function in the presence of past year SO_2_ exposure. SNP rs58475486 is located in a ccRE (SCREEN accession EH37E0279296) and a FOXA1 binding site in the A549 lung adenocarcinoma cell line. FOXA1 has a known compensatory role with FOXA2 during lung morphogenesis in mice (Wan *et al*. 2005). Deletion of both FOXA1 and FOXA2 inhibited cell proliferation, epithelial cell differentiation, and branching morphogenesis in fetal lung tissue. Further functional validation on the effect of rs58475486 on binding affinity of FOXA1 is necessary to confirm whether the role of FOXA1 in this ccRE is important for *KITLG* regulatory and lung function.

The higher frequency of the protective alleles of both rs73440122 and rs58465486 in African populations appears to contradict previous findings that African ancestry was associated with lower lung function (Kumar *et al*. 2010). One possible explanation for this seeming inconsistency is that FEV_1_ is a complex trait whose variation is influenced by many genetic variants of small to moderate effect sizes whose influence on lung function may vary by exposure to environmental factors. We found suggestive evidence that the interaction between rs73440122 and first year exposure to SO_2_ reverses the positive association of rs73440122 with lung function to a negative one (Table 3). When assessed independently, our genetic association analysis showed that the protective A allele of rs73440122 was associated with higher lung function. However, with increasing levels of SO_2_ exposure in the first year of life, increasing copies of the A allele of rs73440122 were associated with decreased lung function. Air pollution is known to negatively impact lung function, and we have previously shown that the deleterious effects of air pollution on lung phenotypes may be significantly increased in African American children compared to other populations experiencing the same amount of exposure (Nishimura *et al*. 2013). It has also been reported that Latino and African American populations often live in neighborhoods with high levels of air pollution (Mott 1995). The increased susceptibility to negative pulmonary effects from air pollution exposure coupled with the disproportionate exposure to air pollution experienced by the African American population may also contribute to the lower lung function seen in this population despite the presence of protective alleles. The overlap of the SPI1 binding site with rs73440122 further supports gene-by-SO_2_ interaction at this locus, since SPI1 played a critical role in the development of type 2 inflammation in the airways through macrophage polarization (Qian *et al*. 2015). We noted that the rs73440122 A allele also showed an interaction approaching suggestive threshold with past year exposure to SO_2_ that was positively associated with FEV_1_. The difference is not surprising because age of exposure may significantly impact the effect of air pollution on lung function (Reviewed in Usemann *et al*. 2019). Further studies are required to better understand the effect of this suggestive interaction on lung function.

One strength of this study is the interrogation of independent lung function associated signals at our novel locus. We identified evidence of three independent signals: the replicated signal that showed evidence of regulatory functions (an open chromatin region with a SPI1/PU.1 binding site), one signal that showed a statistically significant gene-by-SO_2_ interaction on lung function, and one signal that represents to *KITLG* eQTLs in the nasal epithelial cells together with suggestive gene-by-SO_2_ interaction. Our results demonstrated a glimpse of the complicated genetic architecture behind complex traits.

One limitation of this study is that the FEV_1_ genetic association and the eQTL analyses with *KITLG* were performed in different populations due to data availability constraints. Although we did not have RNA-Seq data from lung tissues from our study subjects, we previously demonstrated that there is a high degree of overlap in gene expression profiles between nasal and bronchial epithelial cells (Poole *et al*. 2014). The direction of effect of the association was the same in GALA II Puerto Rican children with asthma but not statistically significant. This may in part due to the significantly lower African Ancestry in Puerto Ricans compared to African Americans.

We replicated 20 of 45 variants in black UK Biobank subjects and observed conflicting “flip-flop” associations in African Americans from the SAPPHIRE and GCPD-A studies. In the past, flip-flop associations were deemed as spurious results. Traditional association testing approach studies the effect of each variant on phenotype independently and increases the chance of flip-flop associations detected between studies. Differences in study design, sampling variation that leads to variation in LD patterns, and lack of consideration of other disease influencing genetic and/or environmental factors are all potential causes of flip-flop associations (Kraft *et al*. 2009; Lin *et al*.2007). Hence, it is not surprising to observe flip-flop associations when gene and environment interactions were detected at our FEV_1_ GWAS locus. It was previously shown that flip-flop associations can occur between and within populations even in the presence of a genuine genetic effect (Kraft *et al*. 2009; Lin *et al*. 2007). Further functional analysis is thus required to validate the relationship between the candidate variants, *KITLG* and FEV_1_. This may include reporter assays to validate potential enhancer or repressor activity and CRISPR-based editing assays to validate the regulatory role of the candidate variants on *KITLG*. Although literature exists describing KIT signaling for lung function in mice (Lindsey *et al*. 2011), additional knockout experiments in a model animal system may be necessary to study how *KITLG* contributed to variation in lung function.

The average concentration of ambient SO_2_ exposure in our participants (Table 1) was lower than the National Ambient Air Quality Standards. It is possible that SO_2_ acted as a surrogate for other unmeasured toxic pollutants emitted from local point sources. Major sources of SO_2_ in San Francisco Bay Area during the recruitment years of 2006 to 2011 include airports, petroleum refineries, gas and oil plants, calcined petroleum coke plants, electric power plants, cement manufacturing factories, chemical plants, and landfills (United States Environmental Protection Agency 2008; United States Environmental Protection Agency 2011). The Environmental Protection Agency’s national emissions inventory data also showed that these facilities emit Volatile Organic Compounds, heavy metals (lead, mercury, chromium, arsenic), formaldehyde, ethyl benzene, acrolein, 1,3-butadiene, 1,4-dichlorobenzene, and tetrachloroethylene into the air along with SO_2_. These chemicals are highly toxic and inhaling even a small amount may contribute to poor lung function. Another possibility is that exposure to SO_2_ captured unmeasured confounding socioeconomic factors.

This study identified a novel protective allele for lung function in African American children with asthma. The protective association with lung function intensified with increased past year exposure to SO_2_. Our findings showcase the complexity of the relationship between genetic and environmental factors impacting variation in FEV_1_, highlights the utility of WGS data for genetic research of complex phenotypes, and underscores the importance of including diverse study populations in our exploration of the genetic architecture underlying lung function.

## Data Availability

Local institutional review boards approved the studies (IRB# 10-02877). All subjects and legal guardians provided written informed consent. TOPMed whole genome sequencing and phenotype data from SAGE II are available on dbGaP under accession number phs000921.v4.p1. Normalized gene count data for KITLG and supplemental materials are available at figshare.

## ACKNOWLEDGEMENTS

The Genes-Environments and Admixture in Latino Americans (GALA II) Study, the Study of African Americans, Asthma, Genes and Environments (SAGE) Study and E.G.B. were supported by the Sandler Family Foundation, the American Asthma Foundation, the RWJF Amos Medical Faculty Development Program, the Harry Wm. and Diana V. Hind Distinguished Professor in Pharmaceutical Sciences II, the National Heart, Lung, and Blood Institute (NHLBI) [R01HL117004, R01HL128439, R01HL135156, X01HL134589]; the National Institute of Environmental Health Sciences [R01ES015794]; the National Institute on Minority Health and Health Disparities (NIMHD) [P60MD006902, R01MD010443], the National Human Genome Research Institute [U01HG009080] and the Tobacco-Related Disease Research Program [24RT-0025]. MJW was supported by the NHLBI [K01HL140218]. JJ and BEH were supported by the NHLBI [R01HL133433, R01HL141992]. KLK was supported by the NHLBI [R01HL135156-S1], the UCSF Bakar Institute, the Gordon and Betty Moore Foundation [GBMF3834], and the Alfred P. Sloan Foundation [2013-10-27] grant to UC Berkeley through the Moore-Sloan Data Science Environment Initiative. ACW was supported by the Eunice Kennedy Shriver National Institute of Child Health and Human Development [1R01HD085993-01].

The SAPPHIRE study was supported by the Fund for Henry Ford Hospital, the American Asthma Foundation, the NHLBI [R01HL118267, R01HL141485, X01HL134589], the National Institute of Allergy and Infectious Diseases [R01AI079139], and the National Institute of Diabetes and Digestive and Kidney Diseases [R01DK113003].

The GCPD-A study was supported by an Institutional award from the Children’s Hospital of Philadelphia and by the NHLBI [X01HL134589].

Part of this research was conducted using the UK Biobank Resource under Application Number 40375. We would like to thank UK Biobank participants and researchers who contributed or collected data.

Whole genome sequencing (WGS) for the Trans-Omics in Precision Medicine (TOPMed) program was supported by the National Heart, Lung and Blood Institute (NHLBI). WGS for “NHLBI TOPMed: Gene-Environment, Admixture and Latino Asthmatics Study” (phs000920) and “NHLBI TOPMed: Study of African Americans, Asthma, Genes and Environments” (phs000921) was performed at the New York Genome Center (3R01HL117004-02S3) and the University of Washington Northwest Genomics Center (HHSN268201600032I). WGS for “NHLBI TOPMed: Study of Asthma Phenotypes & Pharmacogenomic Interactions by Race-Ethnicity” (phs001467) and “Genetics of Complex Pediatric Disorders-Asthma” (phs001661) was performed at the University of Washington Northwest Genomics Center (HHSN268201600032I). Centralized read mapping and genotype calling, along with variant quality metrics and filtering were provided by the TOPMed Informatics Research Center (3R01HL-117626-02S1; contract HHSN268201800002I). Phenotype harmonization, data management, sample-identity QC, and general study coordination were provided by the TOPMed Data Coordinating Center (3R01HL-120393-02S1; contract HHSN268201800001I). We gratefully acknowledge the studies and participants who provided biological samples and data for TOPMed.

WGS of part of GALA II was performed by New York Genome Center under The Centers for Common Disease Genomics of the Genome Sequencing Program (GSP) Grant (UM1 HG008901). The GSP Coordinating Center (U24 HG008956) contributed to cross-program scientific initiatives and provided logistical and general study coordination. GSP is funded by the National Human Genome Research Institute, the National Heart, Lung, and Blood Institute, and the National Eye Institute.

The TOPMed imputation panel was supported by the NHLBI and TOPMed study investigators who contributed data to the reference panel. The panel was constructed and implemented by the TOPMed Informatics Research Center at the University of Michigan (3R01HL-117626-02S1; contract HHSN268201800002I). The TOPMed Data Coordinating Center (3R01HL-120393-02S1; contract HHSN268201800001I) provided additional data management, sample identity checks, and overall program coordination and support. We gratefully acknowledge the studies and participants who provided biological samples and data for TOPMed.

The authors wish to acknowledge the following GALA II and SAGE study collaborators: Shannon Thyne, UCSF; Harold J. Farber, Texas Children’s Hospital; Denise Serebrisky, Jacobi Medical Center; Rajesh Kumar, Lurie Children’s Hospital of Chicago; Emerita Brigino-Buenaventura, Kaiser Permanente; Michael A. LeNoir, Bay Area Pediatrics; Kelley Meade, UCSF Benioff Children’s Hospital, Oakland; William Rodríguez-Cintrón, VA Hospital, Puerto Rico; Pedro C. Ávila, Northwestern University; Jose R. Rodríguez-Santana, Centro de Neumología Pediátrica; Luisa N. Borrell, City University of New York; Adam Davis, UCSF Benioff Children’s Hospital, Oakland; Saunak Sen, University of Tennessee.

The authors acknowledge the families and patients for their participation and thank the numerous health care providers and community clinics for their support and participation in GALA II and SAGE. In particular, the authors thank the recruiters who obtained the data: Duanny Alva, MD; Gaby Ayala-Rodríguez; Lisa Caine, RT; Elizabeth Castellanos; Jaime Colón; Denise DeJesus; Blanca López; Brenda López, MD; Louis Martos; Vivian Medina; Juana Olivo; Mario Peralta; Esther Pomares, MD; Jihan Quraishi; Johanna Rodríguez; Shahdad Saeedi; Dean Soto; and Ana Taveras.

The authors thank María Pino-Yanes for providing feedback on this study and Thomas W Blackwell for providing critical review on this manuscript.

The content is solely the responsibility of the authors and does not necessarily represent the official views of the National Institutes of Health.

